# The Covid-19 epidemic in the UK

**DOI:** 10.1101/2020.06.19.20135517

**Authors:** Giulia Faggio, Franco Peracchi

## Abstract

In this note we present some stylised facts about the Covid-19 epidemic in the UK using daily time-series data published by Public Health England and the UK Department of Health and Social Care. We model the data on confirmed new cases using standard count-data models estimated at the country and national levels using a fixed rolling window to capture changes in the model parameters over time. We then use the most recent estimated models to carry out a simple real-time forecasting exercise aimed at predicting the date when the number of confirmed new cases will be approximately zero.

## 1 Introduction

The purpose of this note is threefold: first, we present some stylised facts about the Covid-19 epidemic in the UK using daily time-series data published by Public Health England (PHE) and the UK Department of Health and Social Care (DHSC). These data represent the main source of quantitative information on Covid-19 for the UK and its devolved administrations – England, Wales, Scotland and Northern Ireland. They include the number of tests, the number of people tested, the number of confirmed positive cases and the number of deaths (both cumulative totals and daily changes).

Second, we model the data on confirmed new cases using standard count-data models (i.e., Poisson and Negative binomial regression models) estimated at the country and national levels using a fixed rolling window to capture changes in the model parameters over time. We then use the most recent estimated models to carry out a simple real-time forecasting exercise aimed at predicting the date when the number of confirmed new cases will be approximately zero.

Third, we aim at contributing to the discussion about the Covid-19 epidemic in the UK. We closely follow the approach used by Peracchi (2020) in analysing the spread of the corona virus in Italy with the hope that the empirical evidence presented here will help in sorting out possible interpretations of the epidemic process and provide a temporal perspective in the case of the UK.

This note proceeds as follows. Section 2 offers a brief presentation of the DHSC/PHE data. Section 3 contains a descriptive analysis of the number of confirmed cases and deaths. Section 4 introduces our basic statistical model. Section 5 presents the results of the forecasting exercise. Section 6 concludes.

## 2 Data

The devolved administrations of England, Wales, Scotland and Northern Ireland have implemented a separate reporting process for Covid-19 data.^1^ As a consequence, data presentation is fragmented and not always provided in a consistent format. Moreover, occasional changes to the national reporting process, as well as repeated revisions of the data series, require additional adjustments. On a daily basis, once national-level data are collected by the devolved administrations, they are sent to the DHSC which combines them into UK-wide figures. Covid-19 data for the country as a whole are then released on the DHSC website each day at about 2:00pm (GMT) and displayed on the PHE dashboard shortly after. Not the same data series are displayed on both websites. The DHSC focuses on UK-wide testing figures; PHE focuses on UK lab-confirmed cases and death counts showing also their breakdown by nation, region and sub-region (or health trust).

Data preparation and release have also been affected by the government attempts to tackle the epidemic. In an effort to boost the number of daily tests, at the start of April 2020 the government pledged to significantly increase the country testing capacity by designing a 5-pillar testing program. Focusing on the first two pillars (which are also the most relevant for our analysis), Pillar 1 refers to swab testing by PHE and National Health Service (NHS) labs; Pillar 2 refers to swab testing delivered by commercial partners (i.e., universities, research centres and private companies) via an expanding network of labs and testing sites across the UK.^2^

When national-level data are separately collected by the devolved administrations, only Pillar 1 related results are reported. When national-level data are combined by the DHSC, the department sums the counts from the four nations and adds data from tests carried out under Pillar 2.^3^ As a result, the UK totals for the number of tests, people tested and lab-confirmed cases do not correspond to the sum of the four nations as Pillar 2 cases are excluded from the national counts.

Conversely, the number of deaths both at the UK and the national levels refers to deaths of people who have had a positive test result confirmed by a PHE or NHS laboratory. Thus, those numbers are based only on confirmed cases detected through Pillar 1 of the government testing program. As a consequence, the UK total of deaths corresponds to the sum of the counts from the four nations. Any discrepancy is largely due to differences in the time national data are collected and reported.

We use daily time-series data on the number of confirmed positive cases and the number of deaths (both cumulative totals and daily changes). These data series are available for the whole of the UK since the start of the outbreak - dated as of 31 January 2020, when the first two confirmed Covid-19 cases in the UK were recorded. In practice, we focus on the period from 28 February 2020 onward, as the first significant daily increase in confirmed cases was recorded on that day. The number of confirmed cases refers to the number of people who tested positive to Covid-19 and whose test result was confirmed by a lab (under both Pillar 1 and Pillar 2 testing schemes). Duplicate tests for the same person are removed and the first specimen date is used as the specimen date for that person. The number of deaths includes deaths of people who have had a positive test result confirmed by a PHE or NHS laboratory (Pillar 1) only. Thus, the death series does not include deaths of people who had Covid-19 but had not been tested, people who were tested positive only via a non-NHS/PHE laboratory, or people who has been tested negative and subsequently caught the virus and died.

## 3 Descriptive statistics

Figure 1 shows the evolution of the daily number of confirmed cases for the UK using three variables: one including cases under both Pillars 1 and 2 (red color); one including cases under Pillar 1 only (green color); and one created by aggregating the counts from the four nations (blue color). For each variable, Figure 1 shows two series: the original series (represented by the thinner profile) and a 7-day centered moving average (represented by the thicker profile). This smoother series is essentially equivalent to taking the weekly changes in the total number of cases divided by seven. By removing both daily noise and day-of-the-week effects, smoothing helps visualizing the time trend.

**Figure 1:**
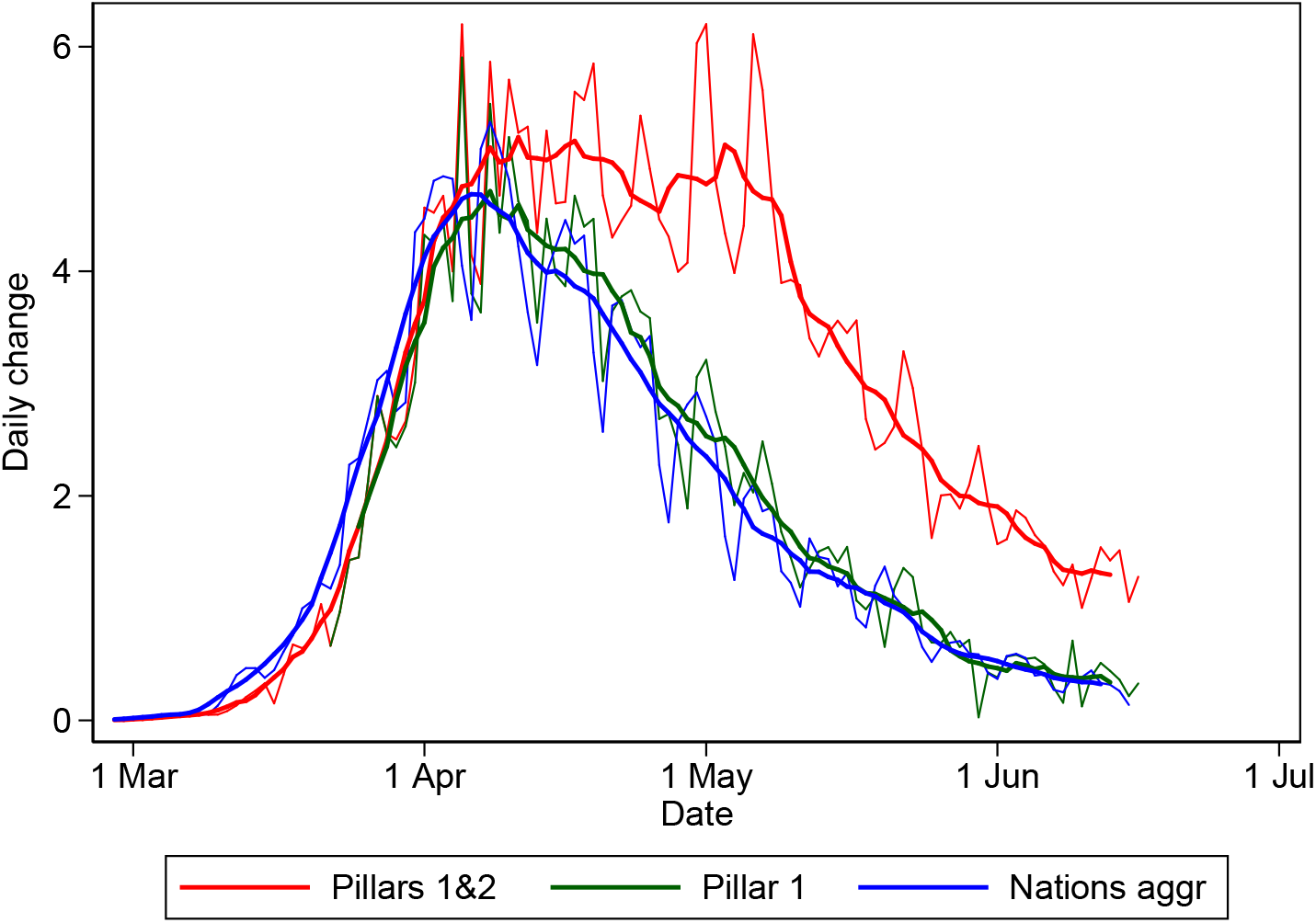
Daily number of confirmed cases (1,000): original series (thinner profiles) and 7-day centered moving averages (thicker profiles).

A data series recording the number of confirmed cases in the UK has been published daily by the DHSC since 25 January 2020. A revised version of the same series starting from 21 March 2020, has been recently (on 28 May 2020) released. After checking that the two series largely coincide over the the period they overlap and that data inconsistencies are restricted to a few episodes, we combine the two series into one. We use the former series up to 21 March 2020, and, from that time onward, we add the running cumulative daily changes from the revised series to the level recorded on 21 March 2020. As a result, we obtain the variable labelled ‘Pillar 1&2’ in Figure 1.

The variable labelled ‘Pillar 1’ is also part of the new (and revised) data series released by the DHSC on 28 May 2020. With the expansion of the government testing program and the creation of its 5-pillar system, there has been an urgency to document the working of each pillar. The variable ‘Pillar 1’ documents the time profile (from 21 March 2020) of confirmed new cases detected via NHS/PHE labs. It is worth noting that variables ‘Pillar 1’ and ‘Pillars 1&2’ coincide till 27 March 2020, when the first five positive cases identified under Pillar 2 were recorded. After 27 May 2020, we observe a surge in the number of confirmed cases detected under the Pillar 2 testing scheme. Both series are falling rapidly since early May.

The variable labelled ‘Nations aggr’ uses the official national counts, also published daily by the devolved administrations – which includes Pillar 1 testing results only. Again, revised versions of these national counts have become available on 28 May 2020. Apart from Scotland, the revised series are somewhat different from the original ones. Still, Figure 1 shows that the official Pillar 1 count (‘Pillar 1’) and the one we have created (‘Nations aggr’) resemble each other quite closely with the main difference being that the variable ‘Nations aggr’ anticipates by a day or two the ‘Pillar 1’ profile. This suggests that there might be a difference in the time of reporting daily figures, with devolved administrations using the date of recording test results and the DHSC using the date of disseminating information. Given the close similarity between the two variables, for the rest of the analysis we will focus on ‘Nations aggr’ to indicate the number of confirmed cases detected under Pillar 1.

Figure 2 shows the time profile of the daily number of confirmed cases (under both Pillar 1 and Pillars 1&2) and the daily number of deaths. As described in section 2, the number of deaths refers to deaths of people who had a positive test result detected through Pillar 1 only; and it includes deaths in all settings, i.e., at hospitals, nursing homes, hospices, etc. Three aspects of Figure 2 are worth highlighting. First, the number of deaths recorded each day (new deaths) are a small fraction of the number of cases confirmed each day under Pillar 1. Second, the profile of new deaths follows with some delay that of the confirmed new Pillar 1 cases and both are hump-shaped. Third, the confirmed new cases under Pillars 1 and 2 combined declines at a smaller rate relative to that of Pillar 1 only. This is true up to the beginning of May, after that the rate accelerates. The first two stylized facts were also reported by Peracchi (2020) in his analysis of Italy. The third is unique to the UK.

**Figure 2:**
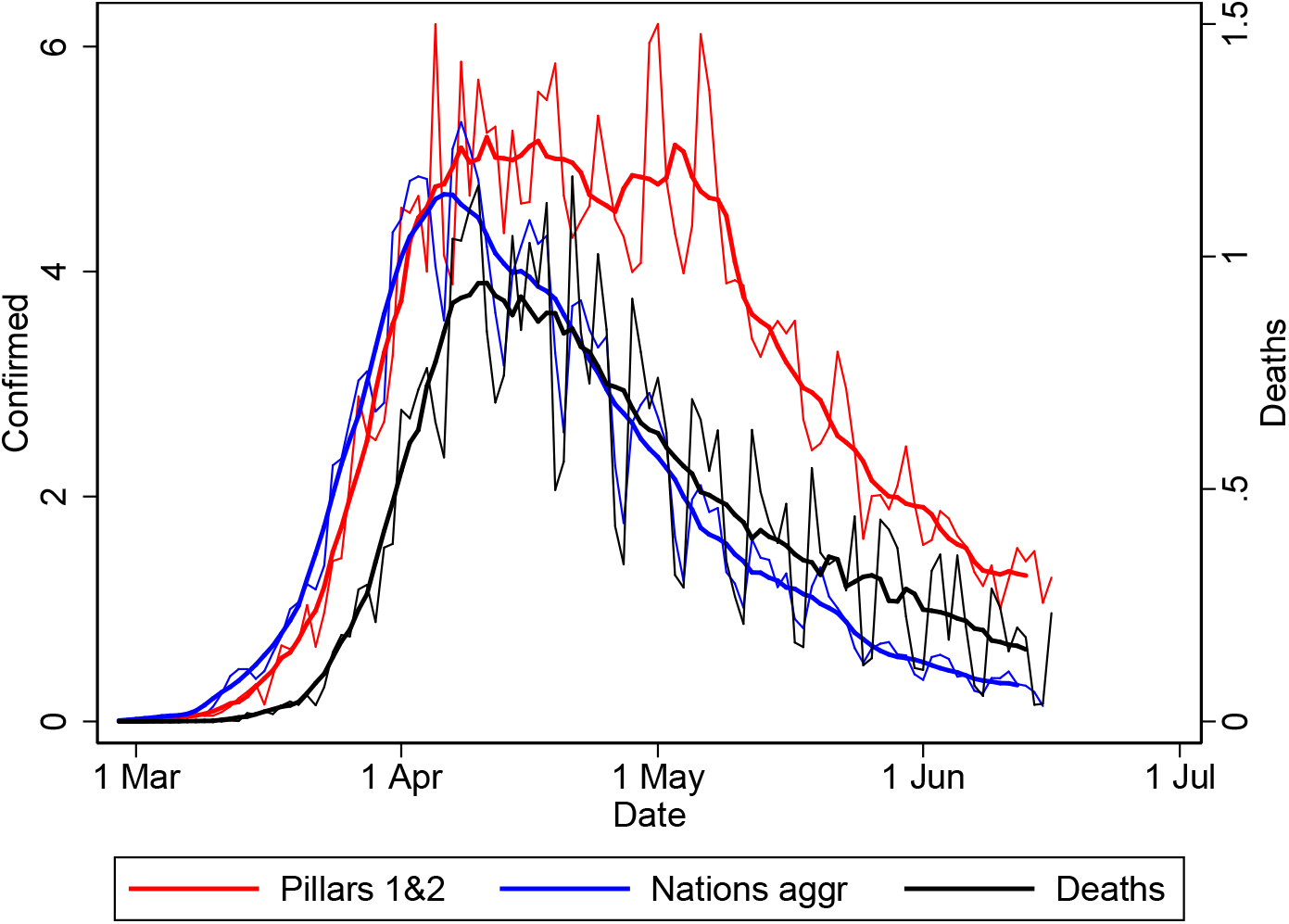
Daily number of new cases (1,000): original series (thinner profiles) and 7-day centered moving averages (thicker profiles). Confirmed cases on left scale, deaths on right scale.

Figure 3 shows the percentage changes in the total number of observed cases for the three variables. The figure reveals a steady downward trend in all series after the very high growth rates till late-March. The time trends observed here are also similar to those observed in other countries.

**Figure 3:**
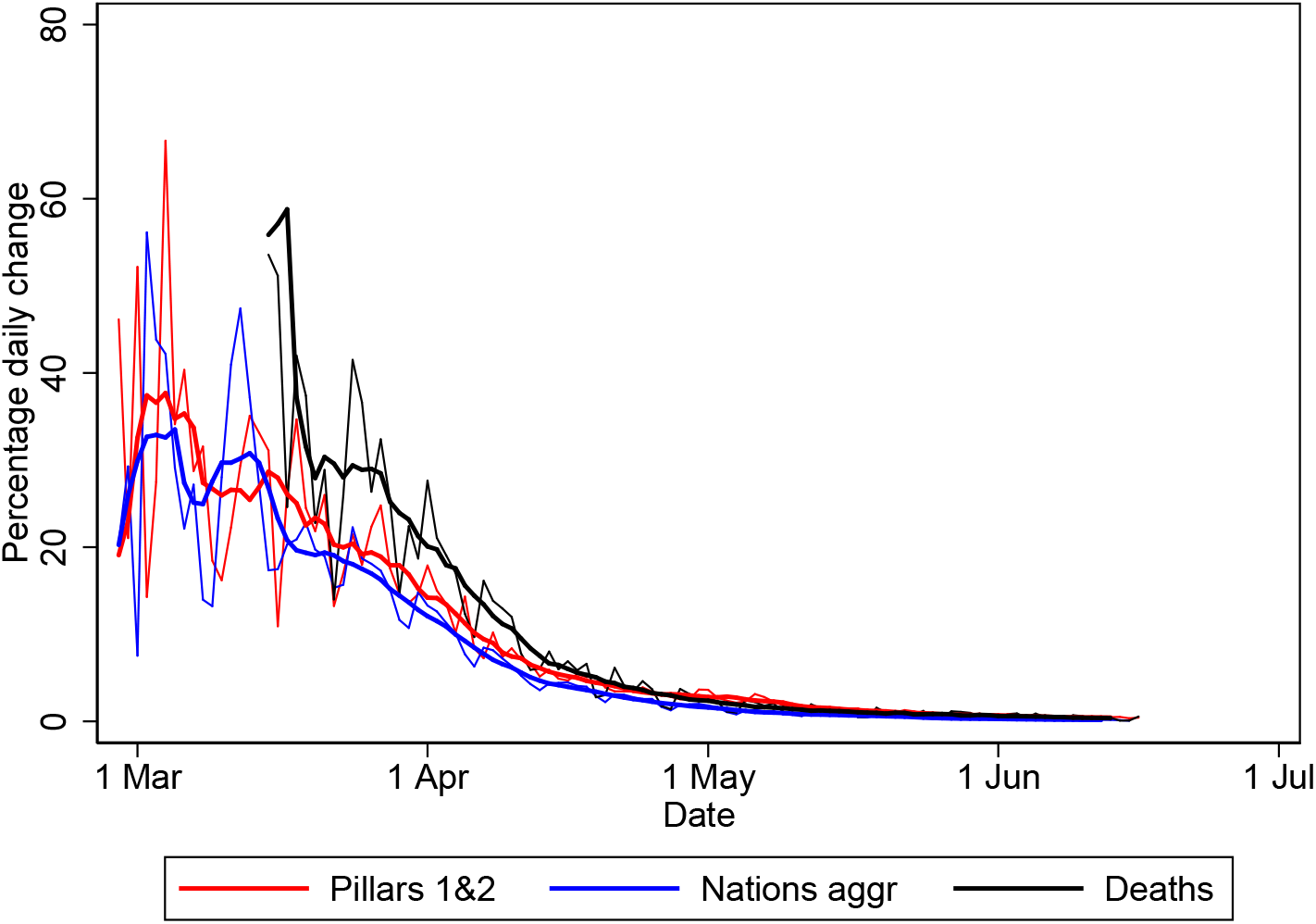
Percentage changes in the number of new cases: original series (thinner profiles) and 7-day centered moving averages (thicker profiles).

Figures 4 and 5 show differences by nation. Apart from the obvious differences in magnitude, the time profile of the plotted series is remarkably similar.

**Figure 4:**
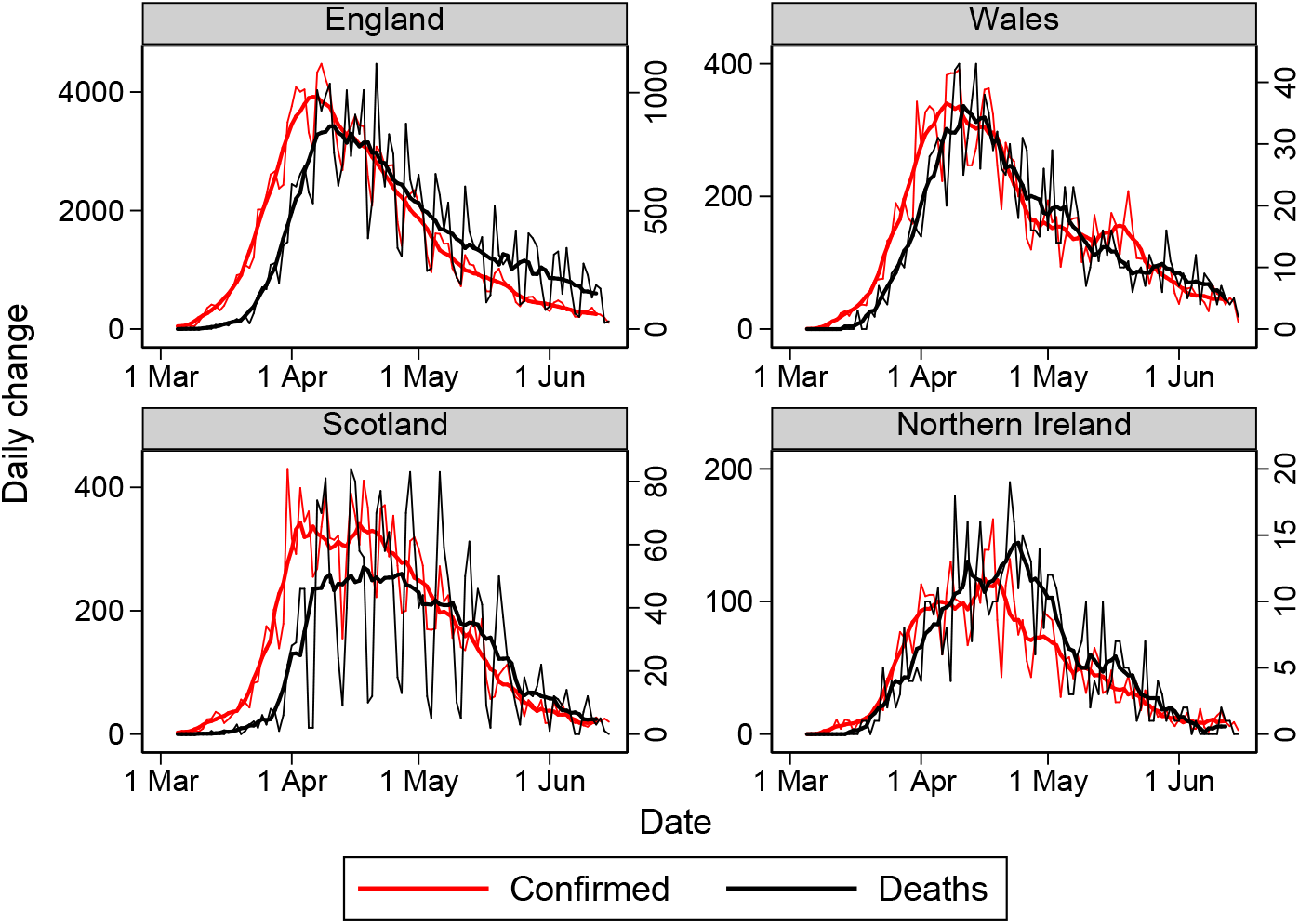
Daily number of new cases by nation: original series (thinner profiles) and 7-day centered moving averages (thicker profiles). Confirmed cases on left scale, deaths on right scale.

**Figure 5:**
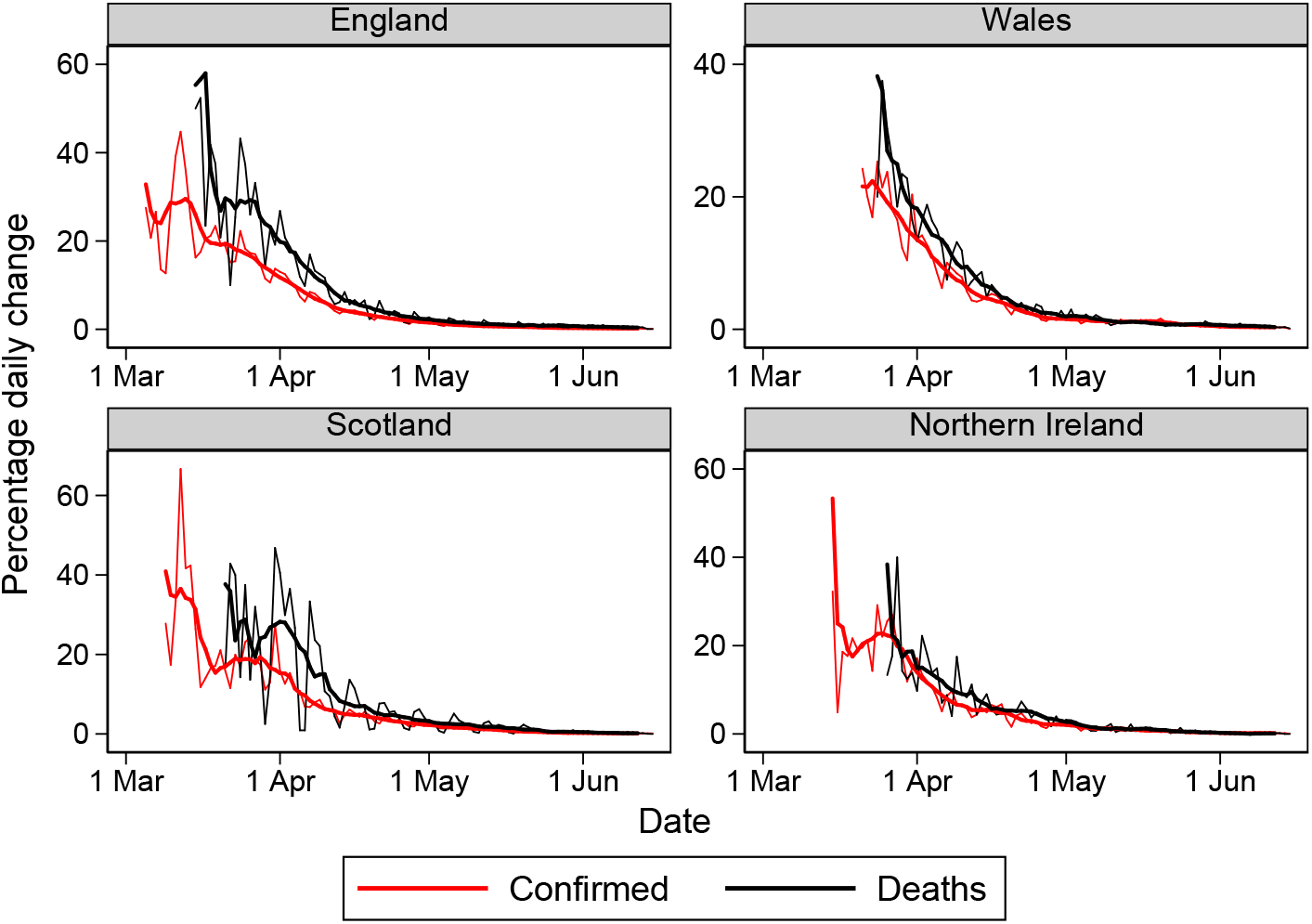
Percentage changes in the number of new cases by nation: original series (thinner profiles) and 7-day centered moving averages (thicker profiles).

## 4 Models

In this section, we focus on modeling the profile of the number of confirmed new cases applying the approach used by Peracchi (2020) in the case of Italy. Specifically, our analysis hinges on the crucial assumption that the daily change in the number of confirmed cases (confirmed new cases) roughly mimics the daily change in the number of newly infected.

We are aware that the total number of confirmed cases is not equal to the unobserved number of people who have ever been infected, but only to the total number of people who have ever tested positive, which is likely to provide a lower estimate of the former for the following reasons. First, eligibility criteria for testing tend to exclude cases that are asymptomatic or not severe enough (see, e.g., Li et al., 2020 and Stock, 2020). Second, available tests may not be fully accurate or their accuracy may be asymmetric, i.e., better at detecting positive cases when truly infected than negative cases when non infected (see Manski and Molinari, 2020). Third, the criteria and the intensity of the testing process are known to vary over time and across space, thereby implying that the unknown ratio between the number of confirmed new cases and the number of the newly infected may also vary. All this suggests that the analysis that follows and the results we obtain should be taken with caution. They might not represent the true underlying process of the pandemic. Still, we believe the exercise is worth pursuing.

Let *Y*_*t*_ denote the number of confirmed new cases. Since *Y*_*t*_ takes nonnegative integer values, a natural model to consider is some parametric count data model. A count data model allows *Y*_*t*_ to be zero, so it would work well both at the beginning and the end of the epidemic, when zero values of *Y*_*t*_ are quite possible.

In this note, we use standard Poisson and Negative binomial (NegBin) models (see, e.g., Cameron and Trivedi, 2013). Both models focus on 𝔼[*Y*_*t*_], the mean value of *Y*_*t*_, which can be very small but nonzero even if the actual value of *Y*_*t*_ is zero at a particular point in time, and both represent this mean as 𝔼[*Y*_*t*_] = exp(*m*_*t*_), where *m*_*t*_ is a deterministic or stochastic function of time, typically modeled as a linear index, that is, a linear combination of time-varying explanatory variables with unknown coefficients. Equivalently, both models represent the natural logarithm of the mean of *Y*_*t*_ as ln(𝔼[*Y*_*t*_]) = *m*_*t*_. The index *m*_*t*_ can be positive or negative, but taking exponentials guarantees that 𝔼[*Y*_*t*_] is strictly positive, though possibly quite small. The main difference between the two models is that while the Poisson model imposes the restriction that the mean of *Y*_*t*_ is equal to its variance 𝕍[*Y*_*t*_], the NegBin model allows for overdispersion, that is, for 𝕍[*Y*_*t*_] *>* 𝔼[*Y*_*t*_]. This is likely to occur when, as in our case, *Y*_*t*_ is obtained by aggregating many heterogeneous units (health trusts or regions of a nation; nations of a kingdom).

For *m*_*t*_, the systematic part of the model, we assume the polynomial form

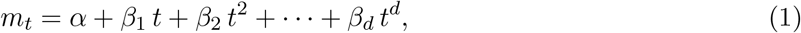

where *α, β*_1_, *β*_2_, …, *β*_*d*_ are unknown parameters to be estimated from the data. In this note we focus on quadratic (*d* = 2), cubic (*d* = 3), and quartic (*d* = 4) specifications of (1).

We fit the resulting Poisson and NegBin models to the two aggregated UK series discussed in Section 2 (Pillars 1&2 and Nations aggr) and then, separately, to the data from each nation, thus allowing for complete heterogeneity at the national level. Estimates in this note use a fixed rolling window to capture possible drifts in the model parameters and thereby improve forecasting. Estimation is carried out by the method of maximum likelihood. Because the sample log-likelihood is strictly concave for both models, just a few iterations are usually enough to locate a global maximum.^4^

Estimates of the various specifications of the two basic models – Poisson and NegBin – are presented in Tables 1 and 2 for the number of confirmed new cases detected under Pillars 1&2 and under Pillar 1 only, respectively. Both tables present results for the UK as a whole. After some experimentation, we select a rolling window of 87 days for both series. Since our estimation period starts on 28 February 2020, the first rolling window of 87 days ends on 24 May 2020. As of 16 June 2020, we have 25 such windows. The estimates in Tables 1 and 2 refer to the most recent window. Standard errors (in parentheses) are robust to heteroskedasticity of unknown form.

**Table 1:** Estimated Poisson and NegBin models for the number of confirmed new cases (Pillars 1&2) in the UK from the most recent window of 87 days ending on 16 June, 2020.

|  | Quadratic trend |  | Cubic trend |  | Quartic trend |  |
| --- | --- | --- | --- | --- | --- | --- |
|  | Poisson | NegBin | Poisson | NegBin | Poisson | NegBin |
| $t$ | -0.0735<br>(0.0000) | -0.0695<br>(0.0000) | -0.0122<br>(0.1132) | -0.0077<br>(0.3089) | -0.0533<br>(0.0024) | -0.0453<br>(0.0025) |
| $t^2$ | -0.0007<br>(0.0000) | -0.0007<br>(0.0000) | 0.0009<br>(0.0000) | 0.0011<br>(0.0000) | -0.0011<br>(0.2165) | -0.0008<br>(0.2715) |
| $t^3$ | | | 0.0000<br>(0.0000) | 0.0000<br>(0.0000) | -0.0000<br>(0.1417) | -0.0000<br>(0.1291) |
| $t^4$ | | | | | -0.0000<br>(0.0270) | -0.0000<br>(0.0150) |
| Constant | -0.3126<br>(0.0020) | -0.2011<br>(0.0299) | 0.2345<br>(0.0005) | 0.2503<br>(0.0001) | 0.0326<br>(0.7600) | 0.0891<br>(0.3091) |
| $n$ | 87 | 87 | 87 | 87 | 87 | 87 |
| $k$ | 3 | 3 | 4 | 4 | 5 | 5 |
| Deviance | 13.997 | 3.7788 | 9.0954 | 2.0855 | 8.4885 | 1.9052 |

*Note:* Estimated standard errors in parentheses.

**Table 2:** Estimated Poisson and NegBin models for the number of confirmed new cases (Pillar 1) in the UK from the most recent window of 87 days ending on 16 June, 2020.

|  | Quadratic trend |  | Cubic trend |  | Quartic trend |  |
| --- | --- | --- | --- | --- | --- | --- |
|  | Poisson | NegBin | Poisson | NegBin | Poisson | NegBin |
| $t$ | -0.1075<br>(0.0000) | -0.0939<br>(0.0000) | 0.0063<br>(0.6026) | -0.0074<br>(0.5172) | -0.1081<br>(0.0000) | -0.0960<br>(0.0000) |
| $t^2$ | -0.0008<br>(0.0000) | -0.0007<br>(0.0000) | 0.0019<br>(0.0000) | 0.0016<br>(0.0000) | -0.0032<br>(0.0005) | -0.0027<br>(0.0011) |
| $t^3$ | | | 0.0000<br>(0.0000) | 0.0000<br>(0.0000) | -0.0001<br>(0.0000) | -0.0001<br>(0.0000) |
| $t^4$ | | | | | -0.0000<br>(0.0000) | -0.0000<br>(0.0000) |
| Constant | -2.1791<br>(0.0000) | -1.8562<br>(0.0000) | -0.9660<br>(0.0000) | -1.0970<br>(0.0000) | -1.6105<br>(0.0000) | -1.5373<br>(0.0000) |
| $n$ | 87 | 87 | 87 | 87 | 87 | 87 |
| $k$ | 3 | 3 | 4 | 4 | 5 | 5 |
| Deviance | 12.856 | 4.0606 | 6.768 | 2.208 | 5.1762 | 1.678 |

*Note:* Estimated standard errors in parentheses.

## 5 Forecasts

Given a particular model specification, our forecast (out-of-sample prediction) of the number of confirmed new cases at some future date *T* + *j* is

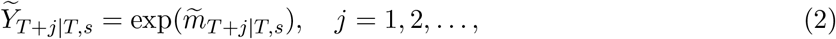

where 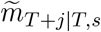 is the predicted value of *m*_*T* +*j*_ based on the estimates from a rolling window of *s* days up to the current date *T*. Thus, 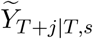 may be interpreted as the expected number of confirmed new cases *j* days ahead given the information contained in the most recent time window of length *s*. When new DHSC/PHE data are released at *T* + 1, we re-run the code on the new data set and produce a new set of forecasts, 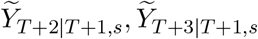 one for each specification considered. This set of forecasts is of interest in itself, and so are the differences from the sets of forecasts obtained at another date.

Given a particular set of forecasts, we can also identify the date when the forecasted number of new cases will approximately be zero. This is an important temporal threshold for decision makers. Because of the forecast rule (2), 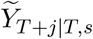 can only be zero in the limit, provided 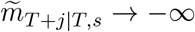 as *j → ∞*. If 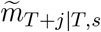 decreases monotonically with *j* and satisfies this condition, then we can take *t*_0_ to be the first date when the forecasted number of confirmed new cases is rounded off to zero (i.e., it takes a value smaller than .5). Notice that *t*_0_ inherits the large statistical uncertainty associated with the whole sequence of forecasts 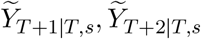 *…*,

All three specifications (quadratic, cubic, and quartic) of the two basic models fit the data reasonably well but they differ considerably in terms of out-of-sample performance. Since we are using a rolling window of fixed length *s*, we can compute (pseudo) out-of-sample forecast errors as the difference 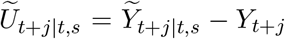 for all *t* + *j ≤ T*. Averaging the squared values of these forecast errors gives an estimate of the mean squared forecast error (MSFE), a standard measure of accuracy of a forecast.

Table 3 shows the estimated MSFE for each model and each specification, separately by devolved administration and for the country as a whole (Pillar 1 and Pillars 1&2 testing schemes). Looking at Pillars 1&2, for both the Poisson and the NegBin models, the quartic specification has the lowest estimated MSFE, followed by the cubic specification. The same is true when considering Pillar 1 only. At the national level, England reports a slightly lower MSFE for the quartic specification than the cubic or the quadratic across both models. Conversely, Wales, Scotland and Northern Ireland report the lowest MSFE for the quadratic specification followed by the cubic and the quartic. This is true for both the Poisson and the NegBin models. In light of these results and since our main objective is to model the two series at the country level (Pillars 1&2 and Pillar 1), the analysis that follows will focus on the quartic specification of both models.

**Table 3:** Mean squared forecast error of the Poisson and NegBin models for different specifications of the index (1).

| Nation | Poisson |  |  | Negbin |  |  |
| --- | --- | --- | --- | --- | --- | --- |
| | $d = 2$ | $d = 3$ | $d = 4$ | $d = 2$ | $d = 3$ | $d = 4$ |
| England | 3.1230 | 3.0764 | 3.0642 | 3.1339 | 3.0788 | 3.0642 |
| Wales | 0.8899 | 0.9457 | 0.9582 | 0.8415 | 0.9266 | 0.9620 |
| Scotland | 0.8719 | 0.9283 | 0.9430 | 0.8252 | 0.9088 | 0.9465 |
| Northern Ireland | 1.0098 | 1.0720 | 1.0874 | 0.9581 | 1.0517 | 1.0913 |
| Pillar 1 | 0.3757 | 0.2008 | 0.1584 | 0.4245 | 0.2159 | 0.1613 |
| Pillars 1&2 | 0.5112 | 0.4036 | 0.3830 | 0.5447 | 0.4318 | 0.4097 |

Our findings are organised as follows: we first present results at the country level using the variable labelled Pillars 1&2. Second, we present our country forecasts based on the variable labelled Pillar 1. Lastly, we describe our findings for the devolved administration.

Focusing on Pillars 1&2, the top panel of Figure 6 shows the predictions, both in-sample and out-of-sample, from the quartic specification of the Poisson and NegBin models estimated from the most recent rolling window. The vertical red line marks the current date (June 17, 2020). The quartic specification seems to fit the actual data quite well and the out-of-sample predictions behave smoothly. The bottom panel of Figure 6 shows the same predictions on the log scale, zooming on the pattern expected when the epidemic is tapering off and is approaching our chosen threshold for the forecasted number of confirmed new cases (.5 cases, or .0005). Notice that pushing this threshold down moves the predicted date *t*_0_ forward in time.

**Figure 6:**
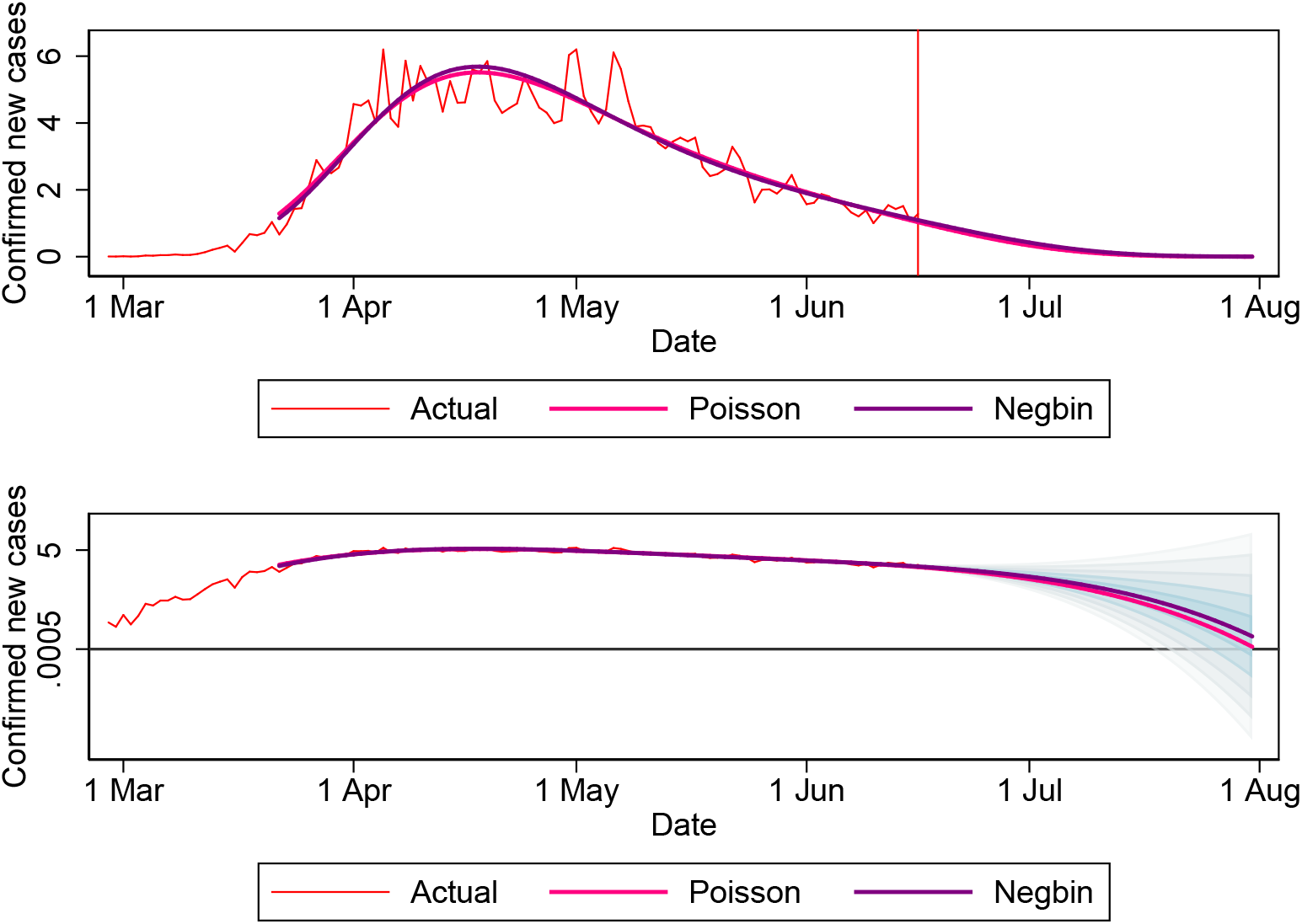
Actual number of confirmed new cases (Pillars 1&2) and current predictions from the quartic specification. *Note*: Top panel shows figures in (1,000); the vertical line marks the current date. Bottom panel shows figures in (1,000, log scale); the vertical red line marks the current date, whereas the horizontal black line marks the threshold for zero confirmed new cases.

The bottom panel of Figure 6 includes “confidence bands” of increasing width for the forecasted values of *Y*_*T* +*j*_, constructed from the estimated parameters of the NegBin model and their standard errors,^5^ and shaded to provide a sense of how forecast uncertainty fans out (for a discussion of “fan charts” of this type see, e.g., Stock and Watson, 2019). These bands are only meant to provide a visual reminder of the large statistical uncertainty associated with this forecasting exercise and cannot be interpreted as a sequence of pointwise confidence intervals on individual forecasts, nor as uniform confidence bands on the whole sequence of forecasts. Notice that the bands for the predicted values of *Y*_*T* +*j*_, constructed vertically and symmetrically at each future date, translate into horizontal asymmetric bands for *t*_0_, the forecasted date of zero confirmed new cases.

The top panel of Figure 7 shows the actual number of confirmed new cases (Pillars 1&2) and current and past predictions from the quartic specification of the NegBin model estimated from various rolling windows. The current (most recent) predictions are in blue, while past predictions are in grey. The vertical red line marks the current date. The bottom panel of Figure 7 shows, on the log scale, the actual number of confirmed new cases and the current and past predictions from the quartic specification of the NegBin model. According to these predictions and our chosen threshold of .5 cases (or .0005), the forecasted number of confirmed new cases will approximately be zero sometime during the second half of July. It is worth noticing that the current prediction is the least optimistic of all.

**Figure 7:**
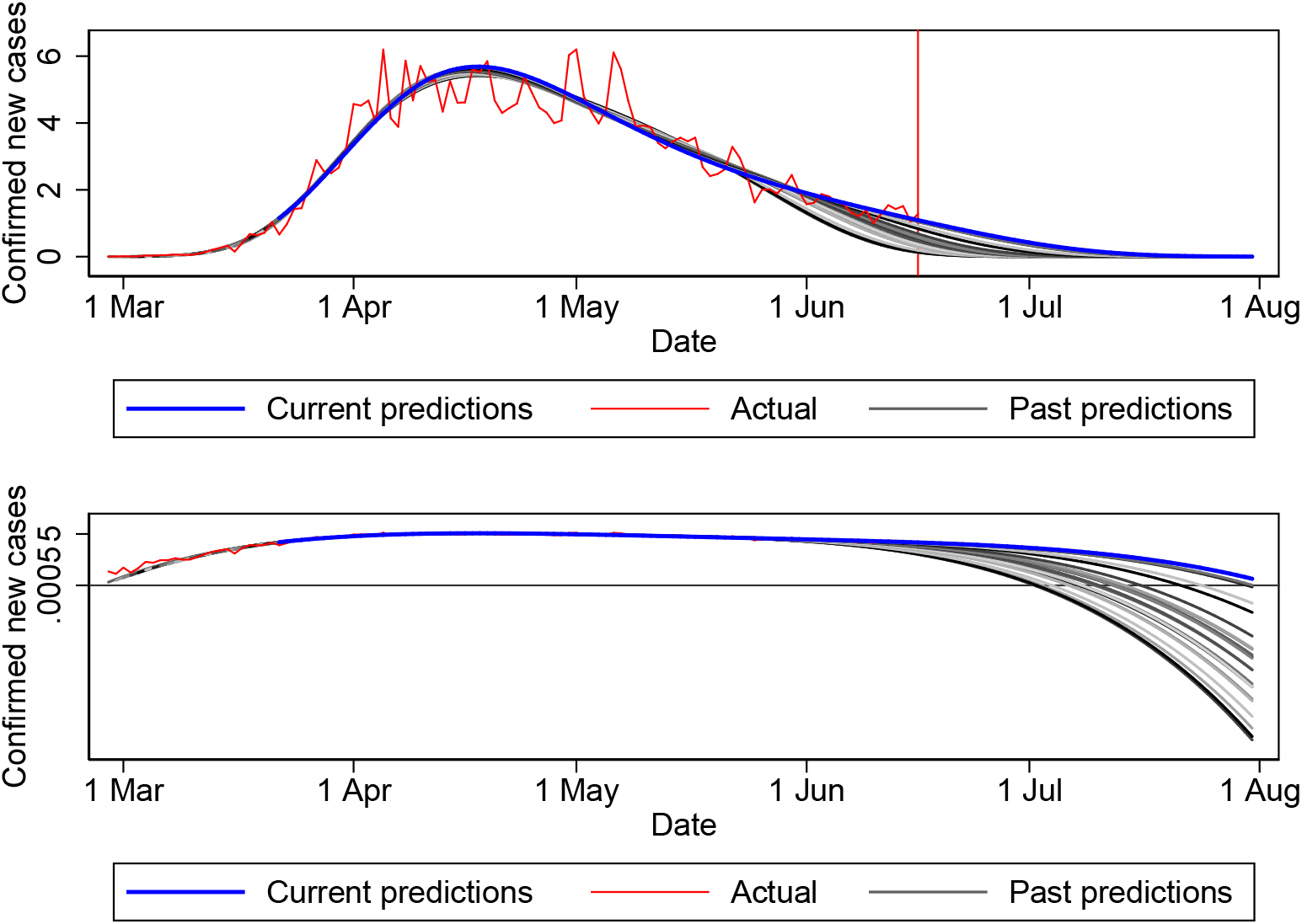
Actual number of confirmed new cases (Pillars 1&2) and current and past predictions from the quartic specification of the NegBin model. *Note*: Top panel shows figures in (1,000); the vertical line marks the current date. Bottom panel shows figures in (1,000, log scale); the vertical red line marks the current date, whereas the horizontal black line marks the threshold for zero confirmed new cases.

To track the accuracy of the out-of-sample forecasts for the UK as a whole (Pillars 1&2), Table 4 presents the actual number of confirmed new cases and its forecasted number by the day of the forecast using the quartic specification of the NegBin model. Table 6 (rows 3 and 4) instead shows the forecasted date of zero new cases by the day of the forecast by using both the Poisson and the NegBin models. For both models, the table indicates that the most recent predictions for Pillars 1&2 are moving *t*_0_ slightly back to a later date.

**Table 4:** Actual number of confirmed new cases (Pillars 1&2) and forecasted number by the day of the forecast. NegBin forecasts from the quartic specification.

| Date | Actual number | Forecasted number by day of the forecast |  |  |  |  |  |  |
| --- | --- | --- | --- | --- | --- | --- | --- | --- |
|  |  | June 9 | June 10 | June 11 | June 12 | June 13 | June 14 | June 15 |
| 10 June | 1.003 | 1.094 |  |  |  |  |  |  |
| 11 June | 1.266 | 1.009 | 0.978 |  |  |  |  |  |
| 12 June | 1.541 | 0.926 | 0.892 | 0.979 |  |  |  |  |
| 13 June | 1.425 | 0.846 | 0.810 | 0.901 | 1.037 |  |  |  |
| 14 June | 1.514 | 0.768 | 0.730 | 0.826 | 0.969 | 1.050 |  |  |
| 15 June | 1.056 | 0.693 | 0.654 | 0.753 | 0.902 | 0.987 | 1.100 |  |
| 16 June | 1.279 | 0.622 | 0.582 | 0.683 | 0.836 | 0.924 | 1.045 | 1.043 |

Source: Figures are expressed in (1,000).

**Table 5:** Actual number of confirmed new cases (Pillar 1) and forecasted number by the day of the forecast. NegBin forecasts from the quartic specification.

| Date | Actual number | Forecasted number by day of the forecast |  |  |  |  |  |  |
| --- | --- | --- | --- | --- | --- | --- | --- | --- |
|  |  | June 9 | June 10 | June 11 | June 12 | June 13 | June 14 | June 15 |
| 10 June | 0.386 | 0.275 |  |  |  |  |  |  |
| 11 June | 0.385 | 0.248 | 0.262 |  |  |  |  |  |
| 12 June | 0.444 | 0.223 | 0.236 | 0.259 |  |  |  |  |
| 13 June | 0.333 | 0.198 | 0.211 | 0.235 | 0.272 |  |  |  |
| 14 June | 0.318 | 0.176 | 0.188 | 0.212 | 0.250 | 0.262 |  |  |
| 15 June | 0.262 | 0.155 | 0.166 | 0.189 | 0.228 | 0.241 | 0.252 |  |
| 16 June | 0.138 | 0.135 | 0.146 | 0.168 | 0.207 | 0.220 | 0.232 | 0.232 |

Source: Figures are expressed in (1,000).

**Table 6:** Forecasted date of zero confirmed new cases by the day of the forecast. Poisson and Negbin forecasts from the quartic specification.

| Data | Model | Forecasted date by day of the forecast |  |  |  |  |  |  |
| --- | --- | --- | --- | --- | --- | --- | --- | --- |
|  |  | June 10 | June 11 | June 12 | June 13 | June 14 | June 15 | June 16 |
| Pillar 1 | Poisson aggr | 6 July | 6 July | 9 July | 10 July | 12 July | 12 July | 11 July |
|  | NegBin aggr | 7 July | 7 July | 13 July | 14 July | 15 July | 15 July | 13 July |
| Pillars 1&2 | Poisson aggr | 11 July | 14 July | 18 July | 21 July | 26 July | 28 July | 31 July |
|  | NegBin aggr | 13 July | 13 July | 22 July | 25 July | 31 July | 31 July | 31 July |

Turning to the variable Pillar 1, Figure 8 shows the actual values of confirmed new cases and the predictions, both in-sample and out-of-sample, from the quartic specification of the Poisson and NegBin models estimated from the most recent rolling window. Figure 8 reproduces for Pillar 1 what Figure 6 did for Pillars 1&2. Similarly to what found in the case of Pillars 1&2, Figure 8 shows that the quartic specification seems to fit the actual data reasonable well and the out-of-sample predictions behave smoothly. Moreover, as in the case of Pillars 1&2, the Poisson and the NegBin models generate out-of-sample predictions that are very close to each other. This finding is also confirmed by Table 6: since 10 June 2020, the two models have been producing estimates of *t*_0_ that are consistently a few days apart from each other.

**Figure 8:**
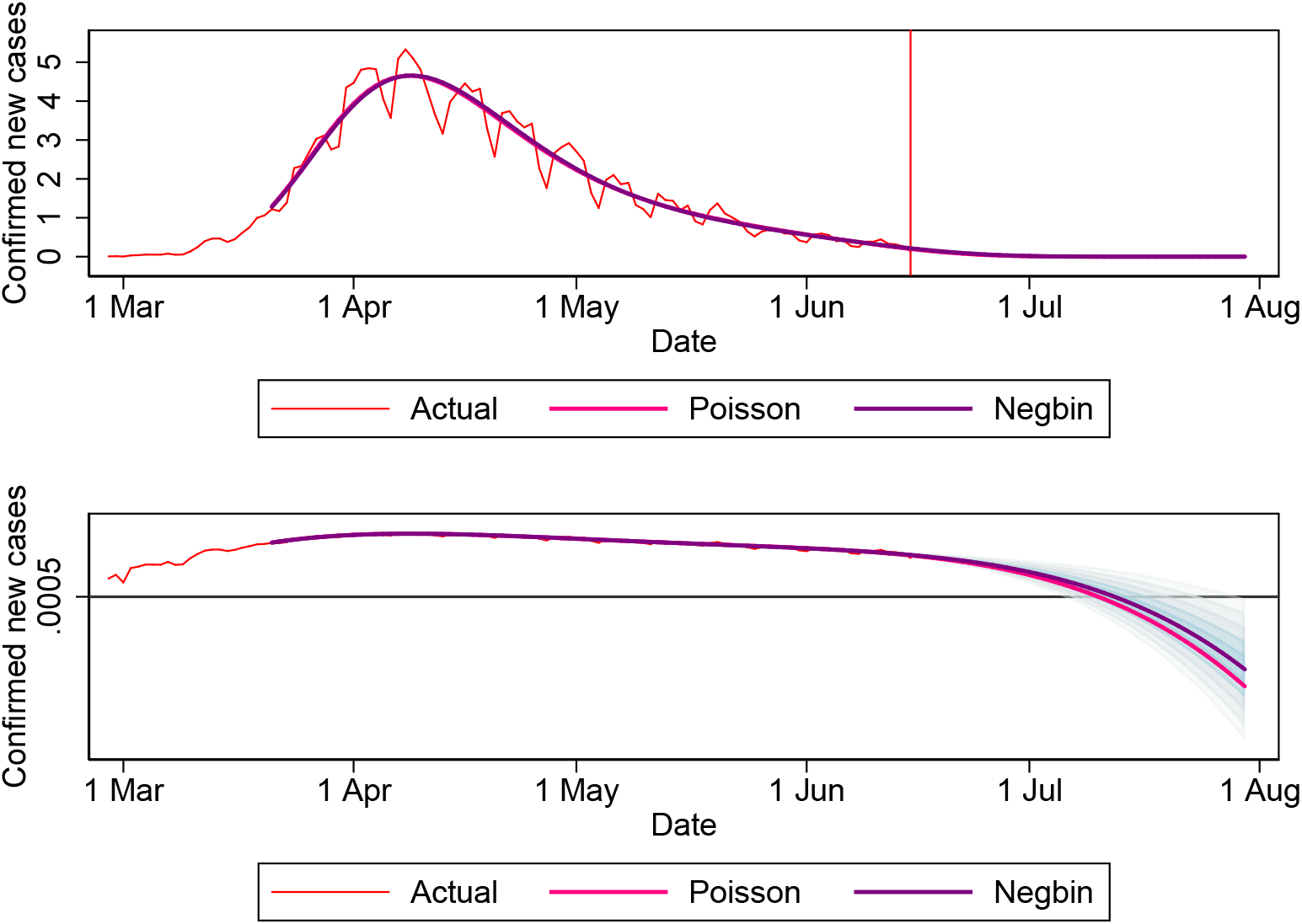
Actual number of confirmed new cases (Pillar 1) and current predictions from the quartic specification. *Note*: Top panel shows figures in (1,000); the vertical line marks the current date. Bottom panel shows figures in (1,000, log scale); the vertical red line marks the current date, whereas the horizontal black line marks the threshold for zero confirmed new cases.

Figure 9 shows actual data as well as current and past predictions from the quartic specification of the NegBin model estimated from various rolling windows. Figure 9 presents for Pillar 1 what Figure 7 presented for Pillars 1&2. Comparing Figure 9 with Figure 7, current predictions are more optimistic than past predictions in the case of Pillar 1; the opposite was true in the case of Pillars 1&2.

**Figure 9:**
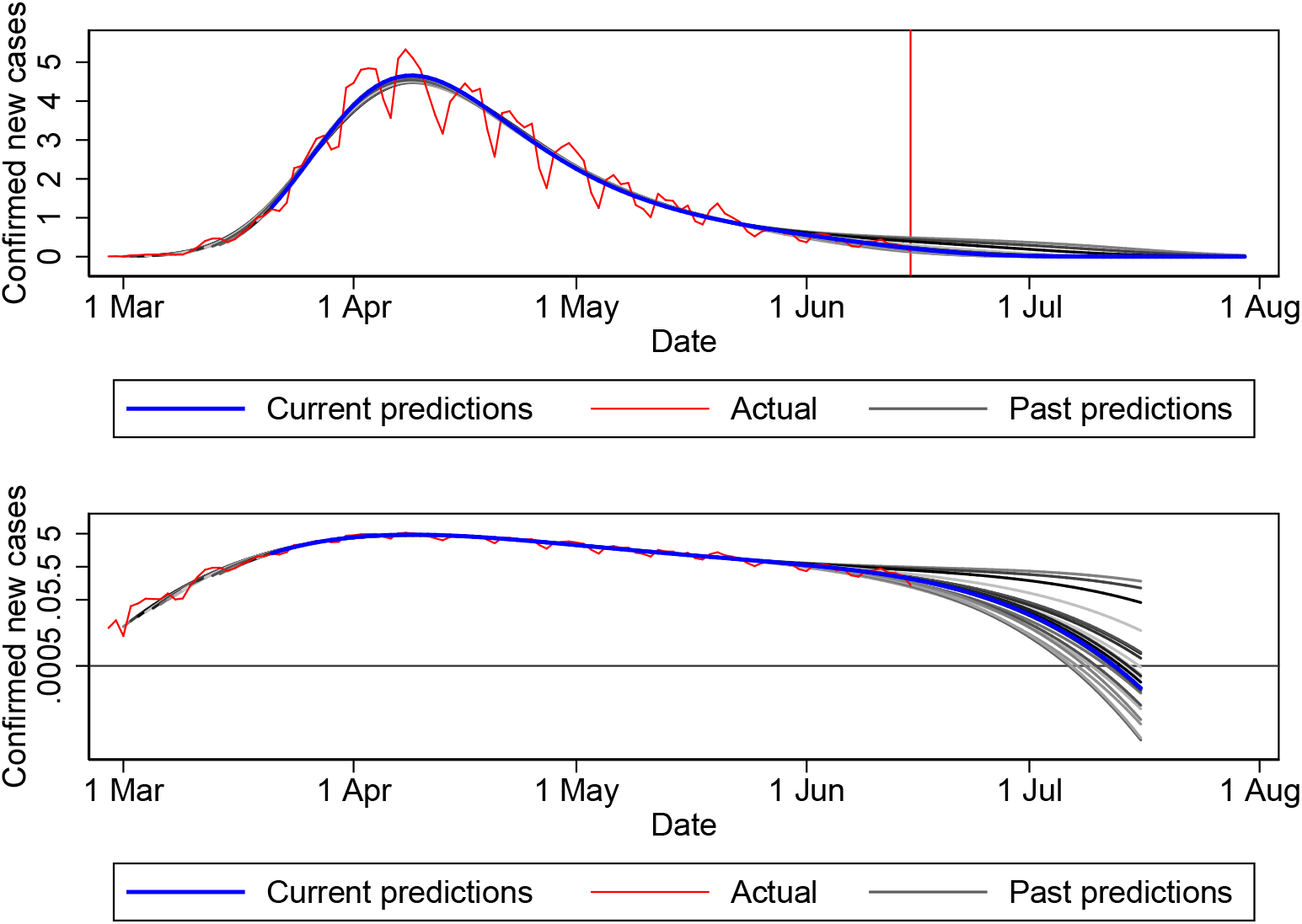
Actual number of confirmed new cases (Pillar 1) and current and past predictions from the quartic specification of the NegBin model. *Note*: Top panel shows figures in (1,000); the vertical line marks the current date. Bottom panel shows figures in (1,000, log scale); the vertical red line marks the current date, whereas the horizontal black line marks the threshold for zero confirmed new cases.

Lastly, we present our findings for the four devolved administrations.^6^ The forecast rule for the number *Y*_*T* +*j,k*_ of confirmed new cases at a future date *T* +*j* in each nation *k* is similar to (2), with the nation-specific parameters estimated using a rolling window of *s* days up to the current date *T*. Again, we choose *s* equal to 87. Figure 10 shows the predictions from the quartic specification of the NegBin model estimated separately by nation. The vertical red lines mark the current date. In creating Figures 10, all past predictions were converging for Scotland and Northern Ireland. Conversely, for England, predictions between 24 May 2020 and 27 May 2020 were dropped because diverging. For Wales, the predictions on 24 May 2020 only were dropped because diverging.

**Figure 10:**
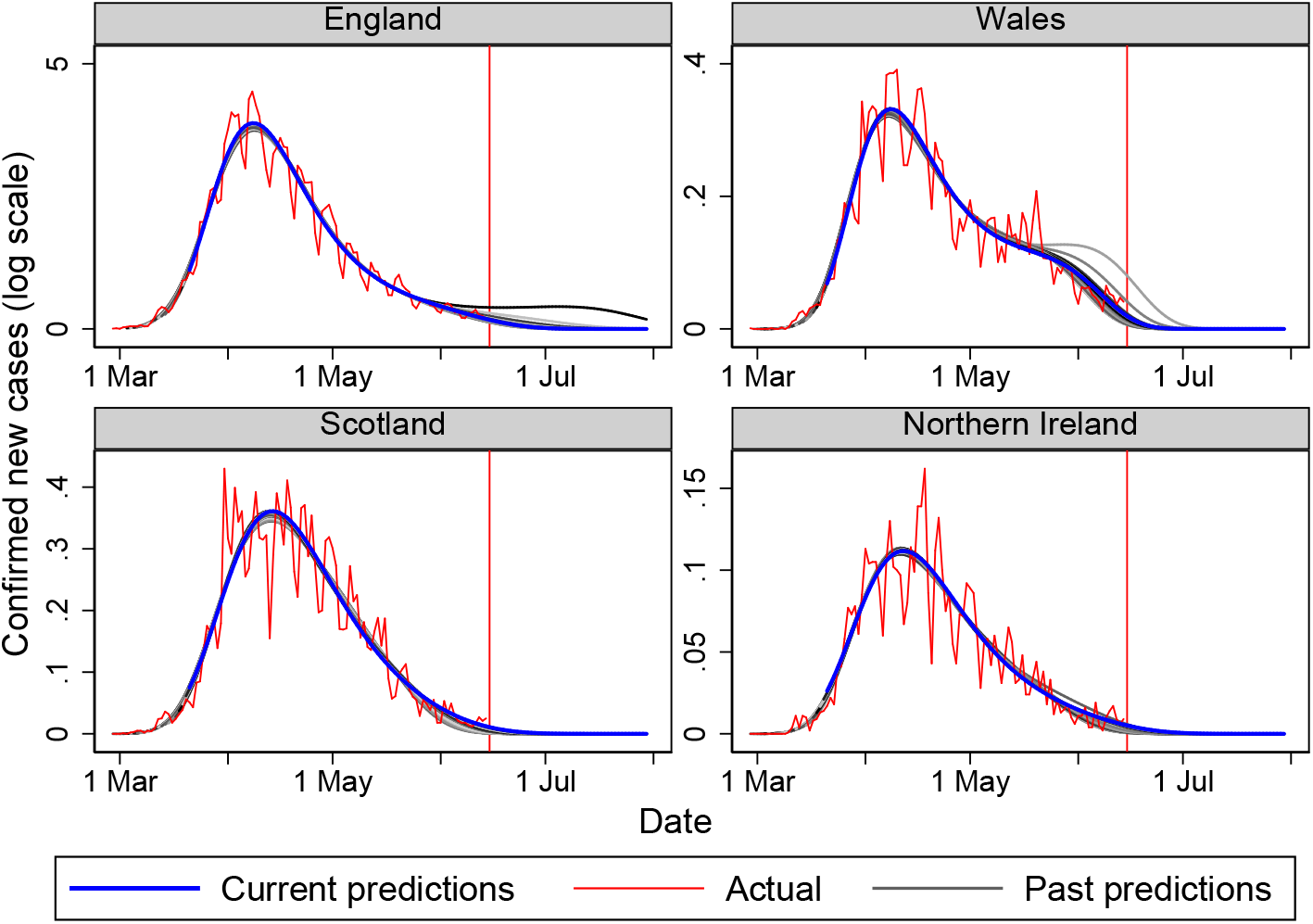
Actual number of confirmed new cases (Pillar 1) by nation and current and past predictions from the quartic specification of the NegBin model. *Note*: Figures are in (1,000). The vertical red lines mark the current date.

Table 7 presents the forecasted date of zero confirmed new cases by the day of the forecast from the quartic specification of the NegBin model estimated separately by nation. According to the latest estimates, all four nations will reach the end of the first phase of the epidemic between late June and early July and within a few days from each other. Past and current predictions suggest that England might be the last nation to reach the end of this first phase.

**Table 7:** Forecasted date of zero confirmed new cases by the day of the forecast. Negbin forecasts from the quartic specification.

| Nation | Forecasted date by day of the forecast |  |  |  |  |  |  |
| --- | --- | --- | --- | --- | --- | --- | --- |
|  | June 10 | June 11 | June 12 | June 13 | June 14 | June 15 | June 16 |
| England | 9 July | 11 July | 15 July | 16 July | 17 July | 15 July | 13 July |
| Wales | 24 June | 25 June | 26 June | 26 June | 28 June | 29 June | 28 June |
| Scotland | 22 June | 22 June | 23 June | 25 June | 27 June | 1 July | 3 July |
| Northern Ireland | 19 June | 22 June | 27 June | 29 June | 29 June | 2 July | 2 July |

Note: Forecasted dates are set to missing (.) when the model produces diverging forecasts.

## 6 Conclusions

After ten weeks of lockdown, on 1 June 2020 the UK government entered the second phase of its strategy to tackle the Covid-19 epidemic by lifting some of the restrictions on economic activity, travelling and social interaction. A further easing of restrictions has been implemented this week (starting on 15 June), although each devolved administration can exercise its territorial jurisdiction on when and how to relax such rules. A key element in guiding the government decision process has been and continues to be the constant monitoring of daily data, including the number of admissions to the hospital due to Covid-19, the number of tests, the number of confirmed new cases, and the number of deaths.

This note has presented some stylized facts on the number of confirmed positive cases and the number of deaths since the outbreak of the epidemic in the UK. It has focused on simple real-time modelling of the time profile of confirmed new cases and on forecasting the date at which their number will be approximately zero. This date has been forecasted for the UK as a whole and separately for each devolved administration.

As for any forecasting exercise, there is a lot of uncertainty surrounding this date. More importantly, ours is a real-time forecasting exercise. As a new data point is added to the data series each day, our forecasting model produces a new forecast, which may vary from yesterday’s one as well as from past forecasts.

The most recent evidence presented in this note suggests that the number of confirmed new cases in the UK is falling at a faster pace than before. Based on this evidence, the forecasted date of zero confirmed cases is expected to fall between the 10th and the end of July, with some devolved administration reaching that target even quicker.

## Data Availability

All the data used in this manuscript are publicly available.

## Footnotes

1 Each administration publishes daily figures on its dashboard drawing information from live systems that are subject to changes and revisions.

2 For more details about the government testing program, see (Department of Health and Social Care, 2020).

3 Public Health Wales is the exception. It collects and reports combined pillars 1 and 2 number of tests, people tested and confirmed cases for Wales. In order to avoid double counting, Pillar 2 numbers for Wales are not added to this final count.

4 To locate the maximum of the sample log-likelihood, we use iteratively reweighted least-squares, which happen to be more stable than the Newton-Raphson method.

5 Because we end up with forecasts very close to the origin, confidence bands constructed using the delta method include negative values. To avoid the problem we construct the bands by inverting the symmetric pointwise standard- error bands centered at 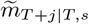

6 There is greater uncertainty surrounding the last five days of national counts than the aggregated variable Pillars 1&2. National counts data are drawn from live systems. Because of this, data from about five days ago can be considered complete; data from recent days are constantly being revised as more information becomes available.

## References

Cameron, A. C. and P. Trivedi (2013). Regression Analysis of Count Data (2nd ed.). New York: Cambridge University Press.

Department of Health and Social Care (2020). Coronavirus (COVID-19) Scaling up our testing pro- grammes. Available at: https://assets.publishing.service.gov.uk/government/uploads/system/uploads/attachmentdata/file/878121/coronavirus-covid-19-testing-strategy.pdf.

Li, R., S. Pei, B. Chen, Y. Song, et al. (2020). Substantial undocumented infection facilitates the rapid dissemination of novel coronavirus (SARS-CoV2). Science (March 16, 2020). Available at: https://science.sciencemag.org/content/early/2020/03/24/science.abb3221.abstract.

Manski, C. F. and F. Molinari (2020). Estimating the COVID-19 infection rate: Anatomy of an inference problem. Journal of Econometrics. Forthcoming.

Peracchi, F. (2020). The COVID-19 pandemic in Italy. Available at: http://www.eief.it/eief/index.php/ forecasts.

Stock, J. H. (2020). Data gaps and the policy response to the novel Coronavirus. NBER Working Paper No. 26902.

Stock, J. H. and M. W. Watson (2019). Introduction to Econometrics (4th ed.). New York: Pearson.

